# Low back pain service utilization and costs: association with timing of first-line services for individuals initially contacting a primary care provider. A retrospective cohort study

**DOI:** 10.1101/2022.06.30.22277102

**Authors:** David Elton, Meng Zhang

## Abstract

**Background:** Low back pain (LBP) is prevalent, costly, and a common reason for a visit to a primary care provider (PCP). Clinical practice guidelines (CPG) recommend a stepped approach to management.

**Objective:** For individuals with LBP initially contacting a PCP, examine the association between the incorporation of first-line therapies, use of second- and third-line services and total episode cost.

**Design:** Retrospective cohort study

**Setting/Patients:** National sample of individuals with non-surgical LBP occurring in 2017-2019.

**Measurements:** Independent variables were initial contact with a PCP, and the timing of incorporation of 5 types of first-line therapies. Dependent measures included use of 13 types of health care services and total episode cost.

**Results:** 8.5% of 118,503 PCPs initially contacted by 308,790 individuals with 347,627 episodes of non-surgical LBP were associated with an episode including any first-line service at any time. 17.5% of episodes included any first-line service at any time. Active care (11.6% of episodes), manual therapy (8.0%) and chiropractic manipulative therapy (7.0%) were the most common first-line services. 5.4% of episodes included a first-line service during the first 7 days with these episodes associated with a modest reduction (risk ratio 0.34 to 0.89) in the use of prescription pharmaceuticals. First-line services were associated with an increase in total episode cost with the smallest increase associated with chiropractic and osteopathic manipulation. Younger individuals from ZIP codes with higher adjusted gross income were more likely to receive a first-line service in the first 7 days of an episode.

**Limitations:** As a retrospective observational analysis of associations there are numerous potential confounders and limitations.

**Conclusions:** Individuals with non-surgical LBP initially contacting a PCP infrequently receive a first-line service, and if received, it is typically in addition to second- and third-line pharmaceutical, imaging, and interventional services. There is an opportunity to improve concordance with the stepped approach to management described in LBP CPGs.

## Introduction

Low back pain (LBP) is common ^1-3^ and costly.^4^ Management of LBP benefits from the availability of high-quality clinical practice guidelines (CPGs) that describe a stepped approach in which services are sequenced into first-, second- and third-line services.^5-7^ In the absence of red flags of serious pathology, LBP CPGs generally emphasize individual self-management, non-pharmaceutical, and non-interventional services as first-line approaches.^5-7^

LBP is a common source of “low-value” care, described as services generating costs without or with minimal beneficial impact on outcomes.^8,9^ Examples of low-value care for LBP include overuse of imaging, interventional procedures, and some prescription pharmaceuticals such as opioids.^10-13^ Management of LBP that is not concordant with CPGs is associated with the risk of LBP transitioning from an acute to a chronic condition.^14^

The initial contact health care provider (HCP) has been used as a method to evaluate variation in service utilization and cost outcomes for LBP.^15,16^ For individuals with LBP, initial contact with a chiropractor (DC), physical therapist (PT), licensed acupuncturist (LAc), or doctor of osteopathy (DO) providing manipulative therapy is associated with first-line services being provided more frequently than second- or third-line services.^17^ More than half of individuals with LBP initially seek treatment from primary care providers (PCP) and physician specialists (PS) ^17^, with LBP the second most common reason for a visit to a PCP.^18^ When initially contacted by an individual with LBP, PCPs and PSs generally incorporate second- and third-line services more frequently than CPG recommended first-line services.^17^ Several barriers to PCP referral for first-line therapies have been identified ^19-24^, and comparisons of early versus late referral for first-line therapies reveal mixed results.^25-34^

The aim of this retrospective, observational study was to examine the association between the timing of incorporation of active care (AC), manual therapy (MT), chiropractic manipulative therapy (CMT), osteopathic manipulative therapy (OMT), or acupuncture (Acu) services, utilization of other healthcare services, and total episode cost for individuals with non-surgical LBP initially contacting a PCP. The hypothesis was that early incorporation of one or more first-line services would be associated with lower rates of second- and third-line service use, and lower total episode cost.

## Methods

### Study design, population, setting and data sources

This is a retrospective cohort study of individuals initially contacting a PCP for non-surgical LBP. An analytic database was created by linking multiple databases. An enrollee database included de-identified enrollment records and administrative claims data for individuals with LBP. De-identified HCP demographic information and professional licensure status was included in an HCP database. ZIP code level population race and ethnicity data was extracted from the US Census Bureau ^35^, adjusted gross income (AGI) data from the Internal Revenue Service ^36^ and socioeconomic status (SES) Area Deprivation Index (ADI) data, from the University of Wisconsin Neighborhood Atlas^®^ database.^37^ With study data being de-identified or a Limited Data Set in compliance with the Health Insurance Portability and Accountability Act and customer requirements, the UnitedHealth Group Office of Human Research Affairs determined that this study was exempt from Institutional Review Board review. The study was conducted and reported based on the Strengthening the Reporting of Observational Studies in Epidemiology (STROBE) guidelines (Supplement – STROBE Checklist).^38^

It was not possible to differentiate whether first-line services were provided by the PCP, accessed by referral from the PCP, accessed by referral from another HCP, or accessed directly by the individual with LBP. The analysis was unable to control for all potential measurable and unmeasurable confounders, and confounders of known measurable confounders. These include: availability of HCPs offering first-line services convenient to an individual’s home, workplace or daily travel routes including public transportation if used, individual preference for specific services or type of HCP including gender or racial concordance, recommendations from family or friends, nuanced clinical complexity not captured in administrative data, anticipated potential out of pocket costs, and appointment availability within a PCP’s and individual’s timing expectations for HCPs meeting these and other criteria.^39^ As an alternative to blurring the line between association and causation through an incomplete adjustment for typical confounders ^40,41^ using potentially inadequate approaches such as propensity score matching ^42^, actual individual demographic and episodic characteristics and associations are reported for the timing of introduction of each of the first-line services analyzed in the study.

### Cohort selection and unit of analysis

The cohort included individuals aged 18 years and older initially contacting a PCP for a complete episode of LBP commencing and ending during the calendar years 2017-2019. This timeframe was selected to follow the release of the American College of Physicians (ACP) LBP CPG ^5^ in 2017 and before the influence of the COVID-19 epidemic on care patterns in early 2020. All individuals had continuous medical and pharmacy insurance coverage during the entire study period.

Episode of care was selected as the unit of analysis. Episodes have been shown to be a valid way to organize administrative claims data associated with a condition.^43^ The Symmetry^®^ Episode Treatment Groups^®^ (ETG^®^) and Episode Risk Groups^®^ (ERG^®^) version 9.5 methodologies and definitions were used to translate administrative claims data into episodes, which have been reported as a valid measurement for comparison of HCPs based on cost of care.^44^ A previous study found a low risk of misclassification bias associated with using episode of care as the unit of analysis.^17^

The analysis included complete episodes defined as having at least 91-day pre- and 61-day post-episode clean periods during which no services were provided by any HCP for any LBP diagnosis. Excluded from the analysis were LBP episodes including a surgical procedure, or associated with diagnoses of malignant and non-malignant neoplasms, fractures and other spinal trauma, infection, congenital deformities and scoliosis, autoimmune disorders, osteoporosis, and advanced arthritis. These exclusions were made to address a potential study limitation of individuals with more complex conditions, or presenting with red flags of potential serious pathology, confounding the analysis of timing of incorporation of first-line non-pharmaceutical and non-interventional services.

### Variables

Data preprocessing, table generation, and initial analyses were performed using Python (*Python Language Reference, Version 3*.*7*.*5*., n.d.). A goodness of fit analysis was conducted using D’Agostino’s K-squared test. Non-normally distributed data are reported using the median and interquartile range (IQR).

The primary independent variables were initial contact with a PCP, and the timing of incorporation of AC, MT, CMT, OMT, or Acu services. Current Procedural Terminology (CPT®) codes were used to identify first-line services. AC – 97110, 97112, 97530. MT – 97140. CMT – 98940 to 98942. OMT – 98925 to 98929. Acu – 97810, 97811, 97813, 97814. For LBP, these are the most frequently provided first-line services recommended by CPGs and covered by commercial insurance.^17^ Passive therapies were excluded from the definition of first-line services. The timing of incorporation of AC, MT, CMT, OMT, or Acu services was based on the number of days after the initial visit with a PCP when a first-line service was first billed by any HCP.

The PCP HCP category consisted of Family Practice, Internal Medicine, General Medicine, and OBGYN physician types, along with Nurse Practitioner and Physician Assistant HCPs. The study cohort was able to access all PCP HCP types directly without a referral. The analyses included a random effect to address variation in decision-making among individual HCPs of the same type.

The primary dependent variable was the rate and timing of use of 13 types of health care services segmented into first-, second-, and third-line service categories based on the ACP LBP CPG as a primary source for the designation ^5^. Secondary dependent variables included the total cost of care for all reimbursed services provided by any HCP during an episode, the number of different HCPs seen during an episode, and episode duration measured in days. Total episode cost included costs associated with all services provided for an episode of LBP, including those not specifically identified in the 13 categories used in the analyses. Costs for services for which an insurance claim was not submitted were not available. The episode duration was the number of days between the first and last date of service for each episode.

Odds (OR), risk (RR) ratios, and associated 95% confidence intervals, were calculated for the timing of introduction of each first-line service type. RR were reported as the measure more widely understood in associational analyses and due to the tendency for ORs to exaggerate risk in situations where an outcome is relatively common.^45^ The RR baseline was episodes where the specific first-line service was not provided. Bivariate analyses were performed comparing episode attributes associated with timing of introduction of AC, MT, CMT, OMT, or Acu services. The bivariate reference baseline was episodes that did not include the specific first-line service. Fisher’s Exact test (p value of .001) was used for comparing the percent of episodes including a service, and Mann Whitney U test (p value of .001) was used for measures reported using median and IQR.

### Role of Funding Source

None

## Results

The sample included 308,790 individuals, with a median age of 47 (Q1 36, Q3 55), and 53.9% females. These individuals were associated with 347,627 complete non-surgical LBP episodes involving 118,503 unique PCPs. There were $208,459,561 in reimbursed health care expenditures with a median total cost per episode of $145 (Q1 $47, Q3 $426). The median pre-episode clean period was 619 days (Q1 403, Q3 853). The median number of days between sequential episodes was 211 (Q1 121, Q3 348). The median post-episode clean period was 432 days (Q1 266, Q3 687) (Table 1). Individuals were from all 50 States and some U.S. territories (Supplement - State).

**Table 1.** Cohort characteristics - % or Median (Q1,Q3)

|  |  |
| --- | --- |
| # of Individuals | 308790 |
| # of Episodes | 347627 |
| # of PCP health care providers (HCPs) | 118503 |
| Total cost | 208459561 |
| Individuals with low back pain |  |
| % Female | 53.9% |
| Age | 47 (36, 55) |
| ERG<sup>®</sup> risk score | 1.5 (0.7, 2.9) |
| Individual home address zip code population attributes |  |
| % non-Hispanic white | 72.1% (50.0%, 85.4%) |
| Area Deprivation Index (ADI) | 48 (31, 65) |
| Household Adjusted Gross Income (AGI) | 61410 (48643, 84035) |
| HCP per 1000 population - DC | 0.21 (0.08, 0.41) |
| HCP per 1000 population - PT | 0.16 (0.03, 0.38) |
| HCP per 1000 population - LAc | 0.00 (0.00, 0.02) |
| Episode attributes |  |
| Total cost | \$145 (47, 426) |
| # of HCP seen | 2 (1, 3) |
| Episode duration - days | 22 (1, 148) |
| Clean period - before initial episode - days | 619 (403, 853) |
| Clean period - between sequential episodes - days | 211 (121, 348) |
| Clean period - after final episode - days | 432 (266, 687) |

82.5% of non-surgical LBP episodes did not include a first-line service at any time during an episode. For the 17.5% of episodes that included any first-line service at any time during an episode, AC (11.5% of episodes), MT (8.0%) and CMT (7.0%) were most common. Individuals were more likely to receive skeletal muscle relaxants (34.1% of episodes), prescription NSAIDs (33.8%), radiography (23.2%), opioids (20.7%) and MRI (8.2%) (Table 2).

**Table 2.** Non-surgical LBP initially contacting PCP - episodic service use by number of days (d) into episode when first line service first incorporated.

|  |  |  |  |  |  |  |  |  |  |  |  |  |  |  |  |
| --- | --- | --- | --- | --- | --- | --- | --- | --- | --- | --- | --- | --- | --- | --- | --- |
| Measure | Episodes |  | First Line |  |  |  |  | Passive Therapy | Second Line |  |  |  | Third Line |  |  |
|  | Count | % | AC | MT | CMT | OMT | Acu |  | Rx - NSAID | Rx - MM Relaxant | Imaging - Radiograph y | Imaging - MRI | Rx-Opioid | Spinal Injection | Imaging - CT |
| Total | 347627 | 100.0% | 11.5% | 8.0% | 7.0% | 1.4% | 0.3% | 5.7% | 33.8% | 34.1% | 23.2% | 8.2% | 20.7% | 4.4% | 1.4% |
| Any First Line Service |  |  |  |  |  |  |  |  |  |  |  |  |  |  |  |
| Not Provided - Reference | 286918 | 82.5% | 0.0% | 0.0% | 0.0% | 0.0% | 0.0% | 0.2% | 33.8% | 34.7% | 20.4% | 6.3% | 20.7% | 3.4% | 1.3% |
| 0-7d | 18700 | 5.4% | 62.1% | 44.8% | 32.2% | 20.1% | 1.6% | 30.0% | 21.5% | 26.5% | 27.1% | 9.1% | 13.2% | 7.0% | 1.1% |
| 8-14d | 6754 | 1.9% | 78.1% | 54.3% | 30.9% | 1.8% | 1.5% | 31.8% | 31.3% | 36.1% | 41.5% | 14.5% | 18.0% | 6.3% | 1.4% |
| 15-28d | 8546 | 2.5% | 74.8% | 52.1% | 34.2% | 2.4% | 1.4% | 31.0% | 34.8% | 34.5% | 42.0% | 18.9% | 20.1% | 7.2% | 1.8% |
| 29-60d | 9716 | 2.8% | 66.9% | 46.6% | 44.3% | 2.4% | 1.4% | 32.8% | 36.9% | 32.7% | 41.4% | 22.5% | 22.6% | 9.3% | 2.0% |
| 61-90d | 5196 | 1.5% | 56.9% | 38.2% | 57.0% | 2.7% | 1.2% | 33.1% | 39.6% | 31.8% | 35.8% | 20.9% | 24.5% | 10.2% | 2.0% |
| >90d | 11797 | 3.4% | 61.7% | 41.0% | 51.9% | 2.3% | 1.7% | 34.9% | 47.2% | 33.2% | 41.6% | 24.7% | 29.6% | 13.4% | 3.0% |
| Active Care (AC) |  |  |  |  |  |  |  |  |  |  |  |  |  |  |  |
| Not Provided - Reference | 307602 | 88.5% | 0.0% | 1.1% | 5.0% | 1.3% | 0.2% | 2.1% | 33.5% | 34.1% | 20.6% | 6.3% | 20.6% | 3.5% | 1.3% |
| 0-7d | 10623 | 3.1% | 100.0% | 61.9% | 20.6% | 2.8% | 0.8% | 33.4% | 24.3% | 28.4% | 32.4% | 11.7% | 13.6% | 8.9% | 1.3% |
| 8-14d | 5249 | 1.5% | 100.0% | 64.2% | 14.9% | 0.6% | 0.9% | 31.6% | 31.0% | 37.2% | 44.1% | 16.1% | 16.6% | 6.7% | 1.4% |
| 15-28d | 6535 | 1.9% | 100.0% | 64.0% | 17.0% | 1.0% | 0.9% | 31.1% | 35.7% | 36.6% | 47.7% | 22.6% | 19.6% | 8.2% | 2.1% |
| 29-60d | 6675 | 1.9% | 100.0% | 61.0% | 23.1% | 1.0% | 0.9% | 33.3% | 39.3% | 36.0% | 49.5% | 29.8% | 23.6% | 12.0% | 2.4% |
| 61-90d | 3029 | 0.9% | 100.0% | 57.1% | 31.9% | 1.0% | 0.7% | 34.4% | 43.7% | 37.1% | 46.5% | 32.0% | 27.5% | 15.0% | 2.7% |
| >90d | 7914 | 2.3% | 100.0% | 56.5% | 32.7% | 1.6% | 1.4% | 37.0% | 50.2% | 37.1% | 48.2% | 33.0% | 32.0% | 17.2% | 3.8% |
| Manual Therapy (MT) |  |  |  |  |  |  |  |  |  |  |  |  |  |  |  |
| Not Provided - Reference | 319792 | 92.0% | 4.9% | 0.0% | 5.8% | 1.4% | 0.2% | 3.0% | 33.6% | 34.1% | 21.6% | 6.9% | 20.6% | 3.8% | 1.3% |
| 0-7d | 6893 | 2.0% | 84.9% | 100.0% | 20.1% | 1.0% | 1.4% | 38.8% | 23.6% | 28.0% | 29.9% | 11.5% | 13.2% | 8.0% | 1.0% |
| 8-14d | 3511 | 1.0% | 92.4% | 100.0% | 14.0% | 0.8% | 1.3% | 34.6% | 31.3% | 37.8% | 41.7% | 16.2% | 17.0% | 7.3% | 1.6% |
| 15-28d | 4720 | 1.4% | 91.8% | 100.0% | 14.2% | 1.0% | 1.0% | 33.8% | 34.2% | 36.0% | 45.4% | 20.7% | 18.1% | 7.8% | 2.1% |
| 29-60d | 4981 | 1.4% | 88.3% | 100.0% | 20.3% | 1.1% | 1.5% | 35.6% | 39.2% | 37.6% | 48.3% | 30.1% | 23.8% | 11.6% | 2.3% |
| 61-90d | 2202 | 0.6% | 85.6% | 100.0% | 27.5% | 1.3% | 1.6% | 37.4% | 44.3% | 37.7% | 46.3% | 32.1% | 26.3% | 15.5% | 2.3% |
| >90d | 5528 | 1.6% | 84.8% | 100.0% | 30.7% | 1.6% | 2.2% | 40.6% | 50.0% | 38.2% | 49.0% | 34.4% | 31.7% | 17.8% | 4.0% |
| Manipulation - Chiropractic (CMT) |  |  |  |  |  |  |  |  |  |  |  |  |  |  |  |
| Not Provided - Reference | 323199 | 93.0% | 9.5% | 6.8% | 0.0% | 1.4% | 0.2% | 3.1% | 33.8% | 34.6% | 22.6% | 8.1% | 20.7% | 4.1% | 1.4% |
| 0-7d | 5378 | 1.5% | 41.7% | 26.3% | 100.0% | 0.6% | 1.0% | 43.7% | 23.4% | 30.8% | 31.5% | 7.4% | 16.3% | 8.7% | 1.2% |
| 8-14d | 2024 | 0.6% | 37.1% | 23.4% | 100.0% | 0.9% | 1.3% | 41.0% | 29.4% | 32.0% | 36.1% | 9.7% | 21.4% | 6.8% | 1.2% |
| 15-28d | 2856 | 0.8% | 35.9% | 22.3% | 100.0% | 0.8% | 1.1% | 40.4% | 32.1% | 29.3% | 29.7% | 10.0% | 21.8% | 5.8% | 1.1% |
| 29-60d | 4401 | 1.3% | 35.2% | 23.2% | 100.0% | 0.7% | 1.2% | 39.9% | 32.4% | 25.8% | 29.6% | 9.5% | 20.2% | 5.4% | 1.2% |
| 61-90d | 3042 | 0.9% | 32.5% | 20.6% | 100.0% | 0.7% | 0.6% | 39.4% | 34.3% | 25.1% | 26.1% | 8.8% | 20.2% | 5.7% | 1.0% |
| >90d | 6727 | 1.9% | 39.1% | 25.1% | 100.0% | 1.1% | 1.4% | 41.8% | 42.7% | 27.3% | 35.4% | 14.5% | 25.0% | 9.4% | 1.7% |
| Manipulation - Osteopathic (OMT) |  |  |  |  |  |  |  |  |  |  |  |  |  |  |  |
| Not Provided - Reference | 342891 | 98.6% | 11.5% | 8.0% | 7.1% | 0.0% | 0.3% | 5.7% | 34.0% | 34.3% | 23.4% | 8.3% | 20.8% | 4.4% | 1.4% |
| 0-7d | 3710 | 1.1% | 11.1% | 4.8% | 3.3% | 100.0% | 0.4% | 6.9% | 11.6% | 15.1% | 9.0% | 3.2% | 7.5% | 2.9% | 0.5% |
| 8-14d | 124 | 0.0% | 16.1% | 10.5% | 6.5% | 100.0% | 1.6% | 8.1% | 25.0% | 37.9% | 33.1% | 8.9% | 16.9% | 8.9% | 0.8% |
| 15-28d | 205 | 0.1% | 17.1% | 9.8% | 7.3% | 100.0% | 0.5% | 11.7% | 27.3% | 32.2% | 22.9% | 9.8% | 18.0% | 10.7% | 0.5% |
| 29-60d | 239 | 0.1% | 16.7% | 12.1% | 3.8% | 100.0% | 1.3% | 9.6% | 31.4% | 34.3% | 25.1% | 12.6% | 27.2% | 14.2% | 2.1% |
| 61-90d | 143 | 0.0% | 18.9% | 7.0% | 5.6% | 100.0% | 1.4% | 7.7% | 28.7% | 34.3% | 22.4% | 12.6% | 26.6% | 9.8% | 2.8% |
| >90d | 315 | 0.1% | 29.2% | 20.0% | 12.4% | 100.0% | 1.6% | 15.2% | 35.6% | 41.3% | 30.8% | 23.2% | 32.4% | 20.3% | 1.3% |
| Acupuncture (Acu) |  |  |  |  |  |  |  |  |  |  |  |  |  |  |  |
| Not Provided - Reference | 346720 | 99.7% | 11.4% | 7.9% | 7.0% | 1.4% | 0.0% | 5.7% | 33.8% | 34.2% | 23.2% | 8.2% | 20.7% | 4.4% | 1.4% |
| 0-7d | 215 | 0.1% | 33.5% | 42.3% | 18.6% | 4.7% | 100.0% | 46.5% | 15.8% | 20.5% | 20.9% | 11.2% | 13.5% | 7.4% | 0.5% |
| 8-14d | 81 | 0.0% | 37.0% | 44.4% | 25.9% | 2.5% | 100.0% | 35.8% | 29.6% | 24.7% | 30.9% | 22.2% | 23.5% | 8.6% | 0.0% |
| 15-28d | 110 | 0.0% | 41.8% | 37.3% | 29.1% | 0.9% | 100.0% | 41.8% | 26.4% | 30.9% | 28.2% | 18.2% | 23.6% | 8.2% | 0.9% |
| 29-60d | 139 | 0.0% | 40.3% | 51.1% | 33.8% | 2.2% | 100.0% | 32.4% | 36.7% | 27.3% | 31.7% | 24.5% | 21.6% | 12.2% | 0.7% |
| 61-90d | 90 | 0.0% | 42.2% | 51.1% | 25.6% | 3.3% | 100.0% | 40.0% | 33.3% | 25.6% | 28.9% | 20.0% | 34.4% | 11.1% | 4.4% |
| >90d | 272 | 0.1% | 50.4% | 49.6% | 41.2% | 2.9% | 100.0% | 39.3% | 44.9% | 32.0% | 33.1% | 25.4% | 27.6% | 15.4% | 2.2% |

Cells with red text denote that service usage was not significantly different from the reference of no first line service (Fisher's Exact p > 0.001)
Cells with black text denote that service usage was significantly different than the reference of no first line service (Fisher's Exact p < 0.001)

Within the first 7 days of an episode 5.4% of episodes included one or more of the five first-line services with AC (3.1% of episodes), MT (2.0%) and CMT (1.5%) being most common. When introduced in the first 7 days, first-line services were generally associated with a reduction in exposure to prescription pharmaceuticals (RR 0.34 to 0.89 depending on the service) and an increase in exposure to radiology, MRI, and spinal injection services. A first-line service introduced 8-14 days into an episode was associated with less significant and generally not clinically meaningful reduction exposure to prescription NSAIDs and opioids (RR 0.72 to 1.13), along with an increase in exposure to spinal imaging and injections. When a first-line service was introduced 15+ days into an episode exposure to prescription pharmaceutical, spinal imaging and spinal injections were higher than if a first-line service was never provided (Table 2)(Supplement – Risk Ratio). The RR for exposure to second- and third-line services based on timing of introduction of AC is illustrated in Figure 1, and CMT in Figure 2.

**Figure 1.**
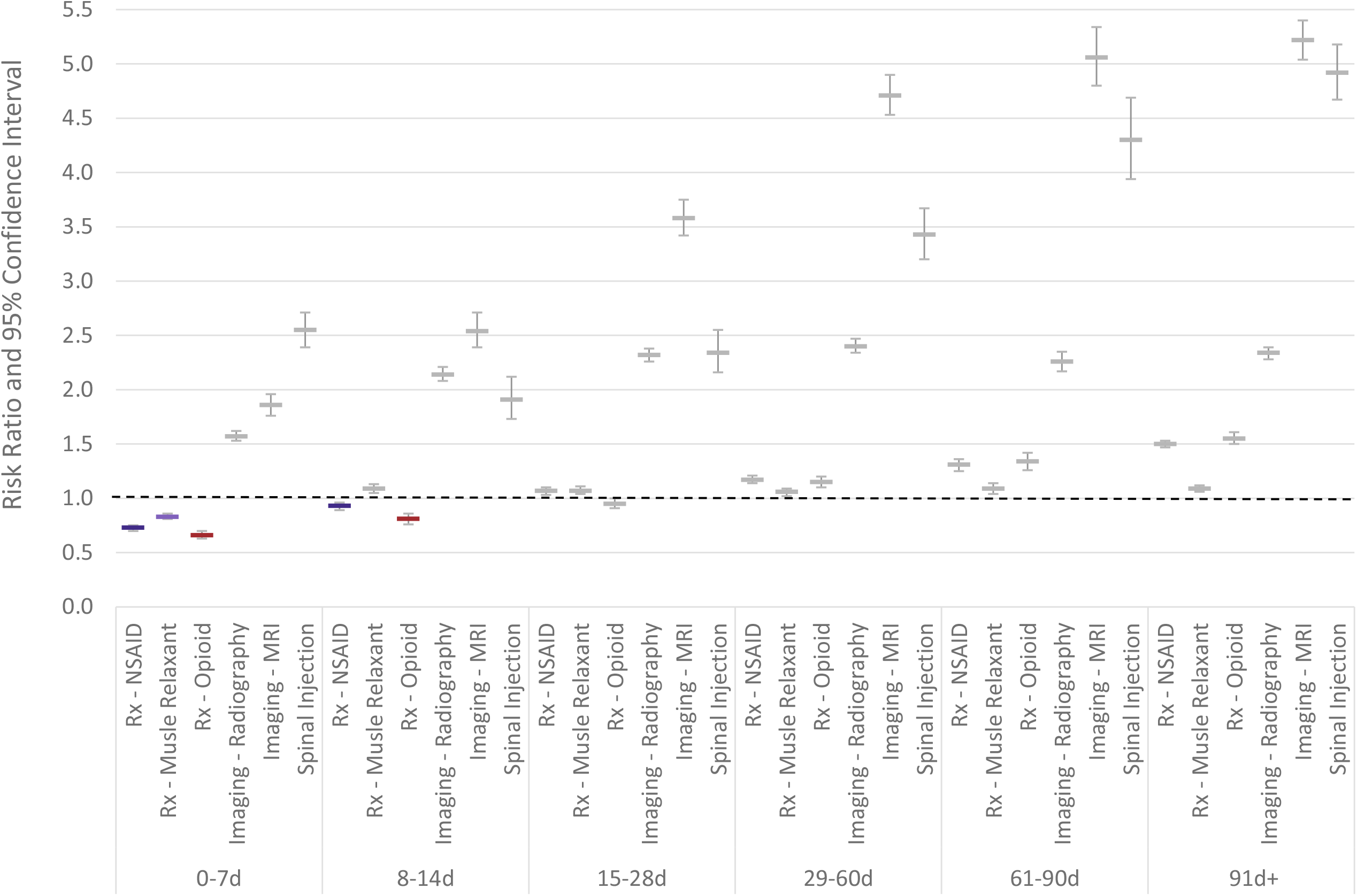
Individuals with non-surgical low back pain initially contacting a primary care provider. Risk ratio and 95% confidence interval for exposure to various health care services based on timing of introduction of **active care** compared to episodes without active care.

**Figure 2.**
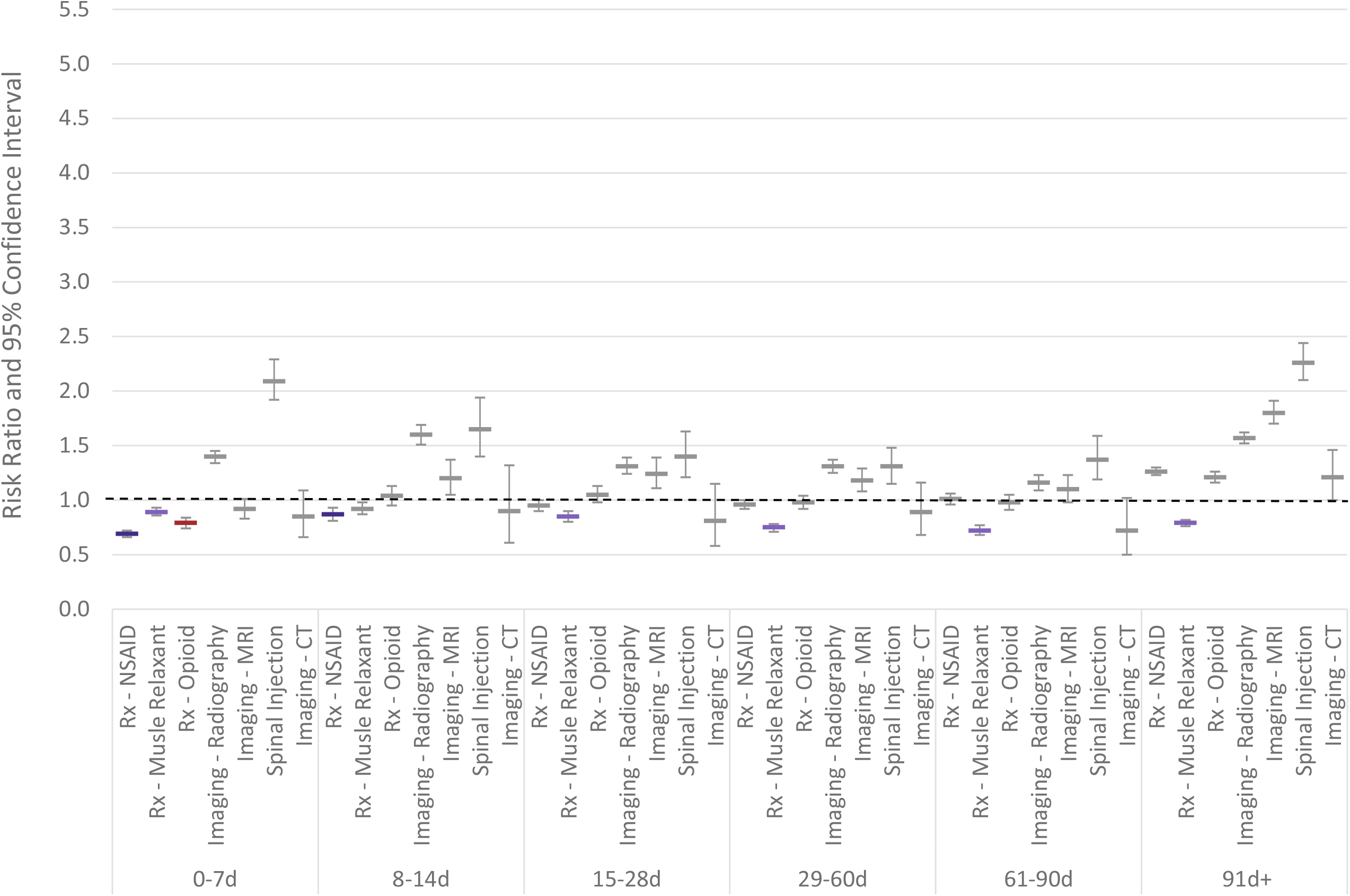
Individuals with non-surgical low back pain initially contacting a primary care provider. Risk ratio and 95% confidence interval for exposure to various health care services based on timing of introduction of **chiropractic manipulative therapy (CMT)** compared to episodes without CMT.

Episodes with a first-line service introduced in the first 7 days were associated with younger individuals, with a lower ERG^®^ risk score, from zip codes with lower deprivation, higher AGI, and greater availability of a DC or PT. Among individual first-line services, Acu was most strongly associated with lower deprivation, higher AGI, lower percent non-Hispanic white population, and greater availability of LAcs (Figure 3)(Table 3).

**Table 3.**
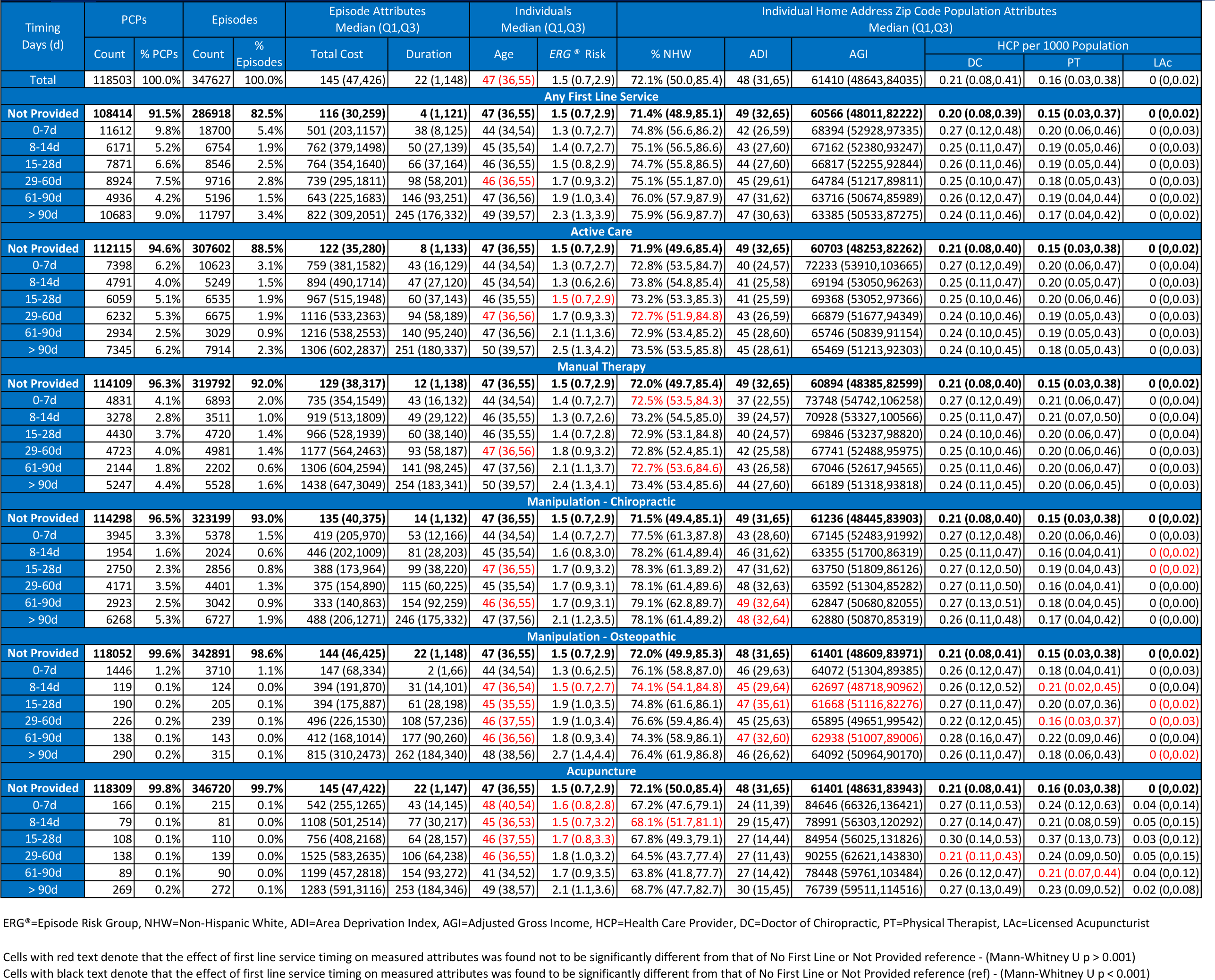
Individual, local population and episode attributes associated with individuals with low back pain initially contacting a Primare Care Provider (PCP) by timing of incorporation of first line services.

**Figure 3.**
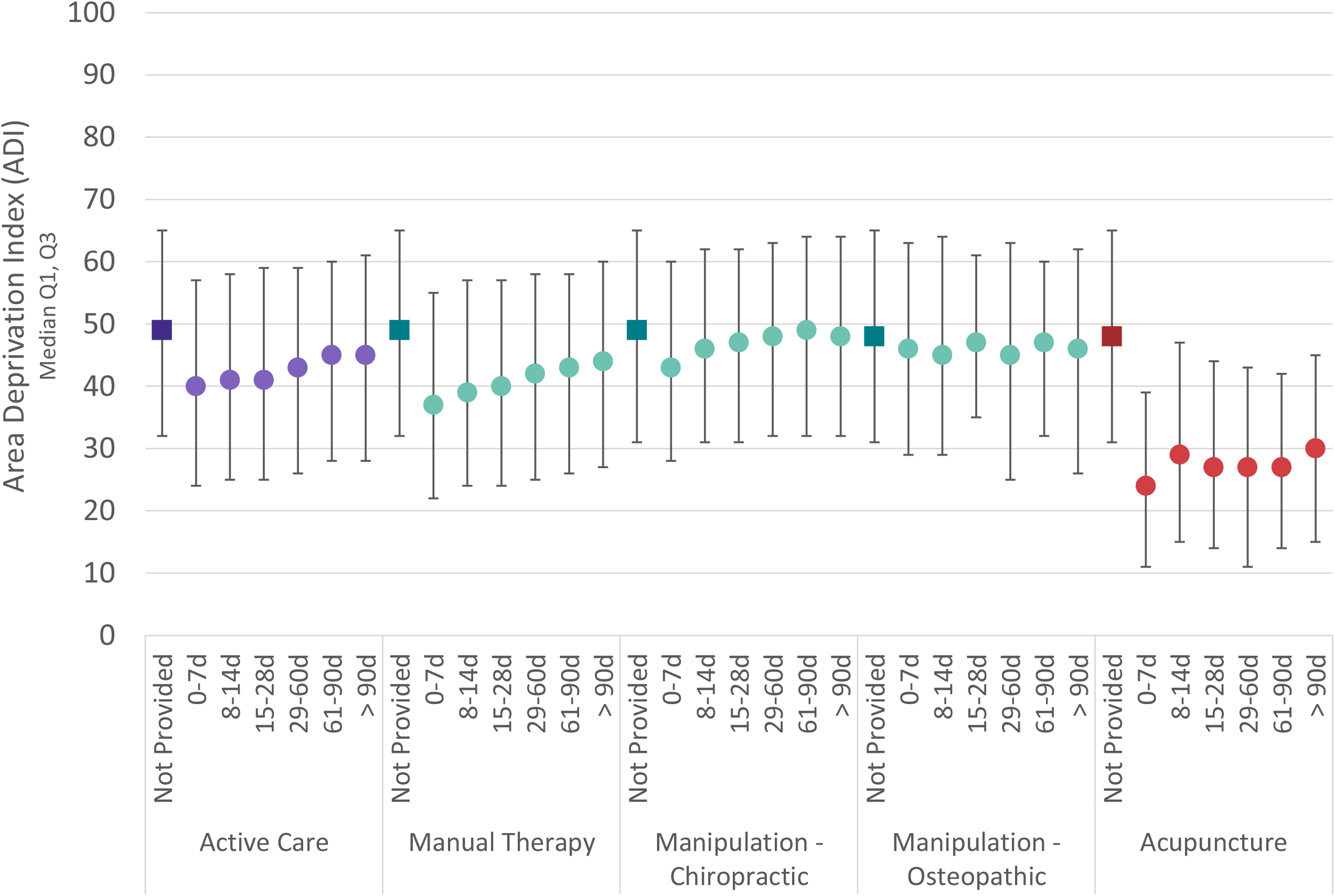
For individuals with low back pain initially contacting a PCP, Area Deprivation Index (ADI) of the individual’s home address zip code associated the number of days (d) into an episode when first line services are initially introduced

Compared to episodes without a first-line service, total episode cost was higher when any first-line service was provided at any time, except for OMT provided within the first 7 days of an episode. The total episode cost increase was lowest for introduction of CMT and OMT (Figure 4). Episode duration increased as first-line services were introduced later in an episode (Table 3).

**Figure 4.**
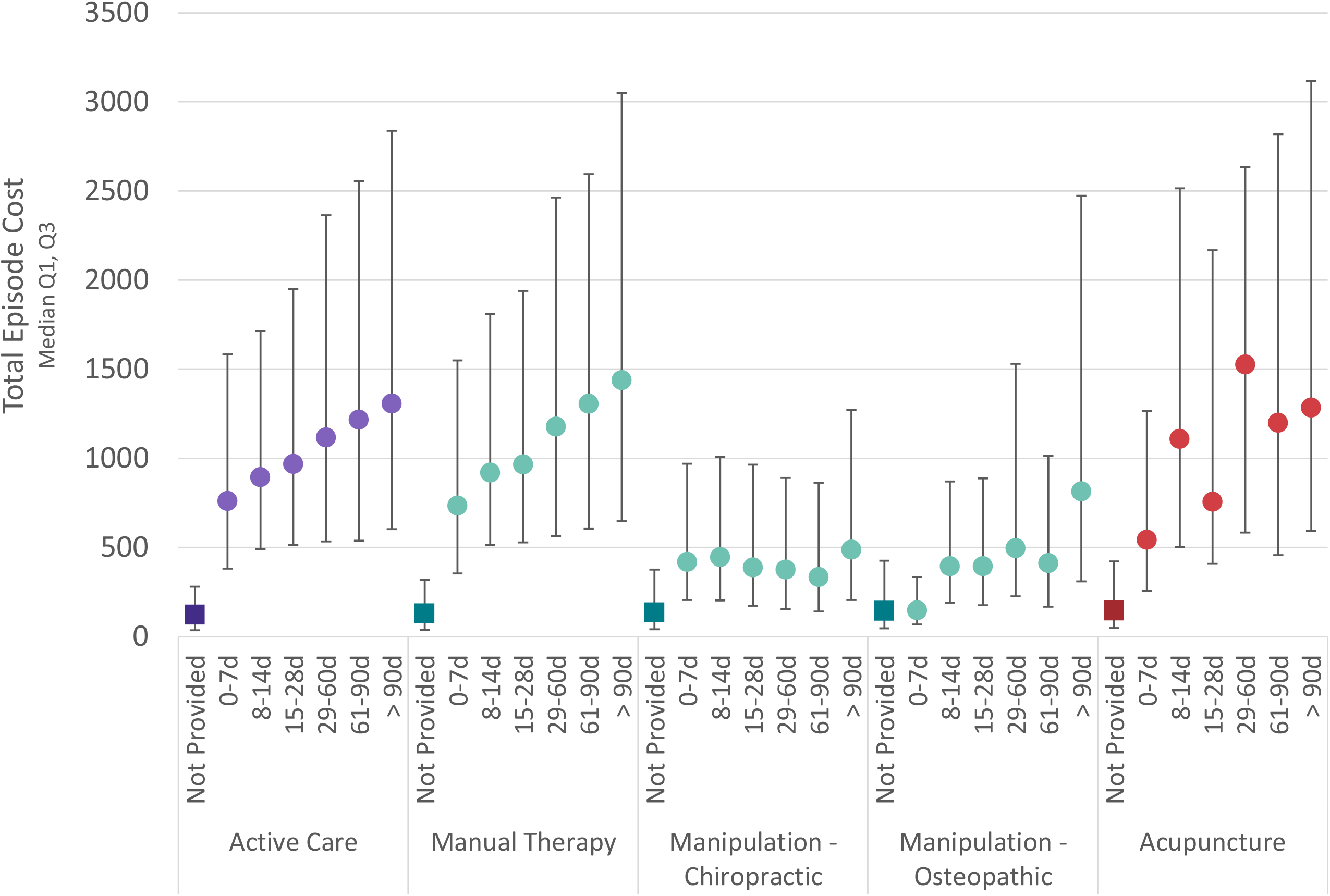
For individuals with low back pain initially contacting a PCP, total episode cost associated with number of days (d) into an episode when first line services are initially introduced

## Discussion

Most individuals with non-surgical LBP initially contact a PCP.^17^ In this study, these individuals received a CPG recommended non-pharmaceutical first-line service at any time in less than 20% of episodes. For the individuals receiving a first-line service within 7 days of initially contacting a PCP there was a modest reduction in exposure to prescription pharmaceuticals. A first-line service introduced more than 7 days after initially contacting a PCP was not associated with a clinically meaningful reduction in any second- or third-line service. Introduction of a first-line service was associated with higher total episode cost, with CMT and OMT associated with the lowest cost increase.

As a retrospective cohort study of associations, several limitations must be kept in mind. The cohort had continuous highly uniform commercial insurance coverage and the processing of administrative claims data included extensive quality and actuarial control measures, nonetheless, data errors, variability in benefit plan design, variability in enrollee cost-sharing responsibility, and missing information were potential sources of confounding or bias. Although the commercial insurer HCP database is under continual validation it may have included errors or missing information regarding the identification of PCPs. Summarizing total episode cost has potential limitations associated with insurance coverage, nature of network participation, and alternative reimbursement models. While individuals from all 50 states and most US territories were included, providing a measure of generalizability, the cohort did not describe a U.S representative sample.

A risk of selection bias was present due to the limited ability to control for individual preference for type of initial contact HCP, individual expectations or requests for specific health care services, and potentially meaningful differences in clinical complexity of individuals with LBP seeking treatment from a PCP. These limitations were partially addressed by excluding LBP episodes associated with significant pathology and by focusing on only non-surgical LBP episodes.

This study expands on an earlier study revealing a low proportion of individuals with LBP initially contacting a PCP have timely incorporation of guideline-adherent non-pharmacologic and non-interventional therapies.^17^ This study corroborates previous studies that found earlier use of first-line therapies may be associated with a reduction in use of low-value services and prescription pharmaceuticals, including opioids.^30,31,46^ The study finding that the benefits of early use of first-line therapies are most evident if initiated within 7 days of initially contacting a PCP corroborates a similar finding ^28^, has potentially important translation implications, and warrants additional study.

Previous studies have found that administrative burden and the cost of non-pharmacologic therapies are perceived as referral barriers by PCPs.^21,22^ This study found first-line therapies were generally incorporated in addition to, rather than as an alternative to, second- and third-line therapies. When used in this way it was not surprising that first-line therapies were associated with higher total episode cost. Incorporating first-line therapies as an alternative to low-value second- and third-line therapies may help address PCP concerns about the cost of non-pharmaceutical therapies. Additional potential barriers to accessing non-pharmacologic health care services that may impact PCP referral may include limited or lack of benefit coverage, wait times for treatment, transportation barriers, the need to secure time away from work to participate in multiple visits, and individual characteristics.^20,47^

Previous studies exploring the association between PCP referral of individuals with LBP for first-line therapies have examined referral for “chiropractic” or “physical therapy”^25,30,31,46^, terms that conflate HCP type and types of health care services that may or may not be provided. This study expands on this earlier work in two ways. First, this study examines the timing of introduction of specific first-line health care services independent of the type of HCP providing the service. Second, this study includes an analysis of the timing of introduction of OMT and Acu services.

## Conclusion

A PCP is commonly the initial HCP consulted by an individual with LBP. High quality CPGs recommend a stepped approach in which favorable natural history, self-care, and non-pharmaceutical services are emphasized as first-line approaches for individuals without red flags of serious pathology. This study revealed second- and third-line pharmaceutical, imaging, and interventional services were commonly provided for individuals with LBP initially contacting a PCP. CPG recommended first-line services were infrequently provided, particularly early in an episode when the potential benefits are greatest. With PCPs frequently consulted for LBP, addressing the high rate of non-concordance with LBP CPGs is important. A variety of individual, local socioeconomic, and HCP availability factors, challenging for a PCP to address in the limited time available during a visit, may present barriers to the timely incorporation of CPG recommended first-line services. A plain language summary of LBP CPGs available to individuals before a visit with a PCP may help improve alignment between CPG recommendations and individual expectations and preferences, making it easier for PCPs to suggest and individuals to follow through on recommendations to incorporate first-line services.

## Supporting information

Supplement - Risk Ratio

Supplement - State Summary

Supplement - STROBE Checklist

## Data Availability

All data produced in the present work are contained in the manuscript

## Declarations

### Ethics approval and consent to participate

Because data was linked from various sources, a review was performed to assess compliance with de-identification requirements. With data being de-identified or a Limited Data Set in compliance with the Health Insurance Portability and Accountability Act and customer requirements, the UnitedHealth Group Office of Human Research Affairs determined that this study was exempt from Institutional Review Board review.

### Consent for publication

Not applicable

### Availability of data and materials

The data are proprietary and are not available for public use but, under certain conditions, may be made available to editors and their approved auditors under a data-use agreement to confirm the findings of the current study.

### Competing interests

At the time of manuscript submission **DE, MZ**, and **PA** are UnitedHealth Group employees. **DE** and **PA** are UNH stockholders. No other potential conflicts of interest or competing interests exist.

### Authors’ contributions

Study conception and design; **DE**. Data acquisition; **DE, MZ**. Data analysis and interpretation; **DE, MZ, PA**. Draft or substantially revise manuscript; **DE, MZ, PA. Registration:** N/A

### Funding Source

None

## List of Abbreviations

LBP: Low back pain
US: United States
CPG: Clinical practice guideline
PCP: Primary care provider
PS: Physician specialist
DC: Doctor of Chiropractic
PT: Physical Therapist
HCP: Health care provider
IQR: Interquartile range
OR: Odds ratio
RR: Risk ratio
AGI: Adjusted Gross Income
ADI: Area Deprivation Index
STROBE: Strengthening the Reporting of Observational Studies in Epidemiology
*CPT*^*®*^: Current Procedural Terminology
*ETG*^*®*^: Episode Treatment Group^*®*^
*ERG*^*®*^: Episode Risk Group^*®*^
ACP: American College of Physicians
PA: Physician Assistant
CMT: Chiropractic manipulative treatment
OMT: Osteopathic manipulative treatment
AC: Active care
MT: Manual therapy
Acu: Acupuncture

## Notes

### Competing Interest Statement

The authors have declared no competing interest.

### Funding Statement

This study did not receive any funding

### Author Declarations

Because data was de-identified or a Limited Data Set in compliance with the Health Insurance Portability and Accountability Act and customer requirements, the UnitedHealth Group Office of Human Research Affairs Institutional Review Board determined that this study was exempt from ethics review

### Summary of Updates

To prepare for submission to peer reviewed journal; minor grammatical updates to manuscript, and updated Figure 3 and 4 labels. No revisions to findings, figures 1/2, tables, supplements or conclusions.

