## Supplement - Risk Ratio for "Low back pain service utilization and costs: association with timing of first-line services for individuals initially contacting a primary care provider. A retrospective cohort study"

Supplement Risk Ratio - Individuals with non-surgical low back pain initially contacting a PCP - risk ratio and 95% confidence interval for service exposure based on timing of introduction of first line services compared to if service not introduced

| | First Line | | | | | Passive Therapy | Second Line | | | | Third Line | | |
|---|---|---|---|---|---|---|---|---|---|---|---|---|---|
| | AC | MT | CMT | OMT | Acu | | Rx - NSAID | Rx - MM Relaxant | Imaging - Radiography | Imaging - MRI | Rx-Opioid | Spinal Injection | Imaging-CT |
| **Active Care (AC)** | | | | | | | | | | | | | |
| 0-7d | N/A | 55.44 (53.46, 57.50) | 4.16 (4.00, 4.33) | 2.08 (1.86, 2.34) | 4.55 (3.61, 5.73) | 15.72 (15.16, 16.29) | 0.73 (0.70, 0.75) | 0.83 (0.81, 0.86) | 1.57 (1.53, 1.62) | 1.86 (1.76, 1.96) | 0.66 (0.63, 0.70) | 2.55 (2.39, 2.71) | 1.02 (0.86, 1.21) |
| 8-14d | N/A | 57.53 (55.33, 59.81) | 3.01 (2.82, 3.22) | 0.48 (0.35, 0.68) | 4.99 (3.69, 6.76) | 14.88 (14.20, 15.58) | 0.93 (0.89, 0.96) | 1.09 (1.05, 1.13) | 2.14 (2.08, 2.21) | 2.54 (2.39, 2.71) | 0.81 (0.76, 0.86) | 1.91 (1.73, 2.12) | 1.15 (0.92, 1.44) |
| 15-28d | N/A | 57.35 (55.22, 59.57) | 3.44 (3.25, 3.63) | 0.78 (0.61, 0.99) | 5.35 (4.10, 6.98) | 14.62 (14.00, 15.27) | 1.07 (1.03, 1.10) | 1.07 (1.04, 1.11) | 2.32 (2.26, 2.38) | 3.58 (3.42, 3.75) | 0.95 (0.91, 1.00) | 2.34 (2.16, 2.55) | 1.64 (1.38, 1.94) |
| 29-60d | N/A | 54.60 (52.55, 56.74) | 4.67 (4.46, 4.89) | 0.75 (0.59, 0.95) | 5.24 (4.01, 6.83) | 15.64 (15.01, 16.31) | 1.17 (1.14, 1.21) | 1.06 (1.02, 1.09) | 2.40 (2.34, 2.47) | 4.71 (4.53, 4.90) | 1.15 (1.10, 1.20) | 3.43 (3.20, 3.67) | 1.90 (1.62, 2.22) |
| 61-90d | N/A | 51.19 (48.92, 53.57) | 6.44 (6.10, 6.79) | 0.77 (0.54, 1.09) | 4.23 (2.77, 6.47) | 16.19 (15.33, 17.10) | 1.31 (1.25, 1.36) | 1.09 (1.04, 1.14) | 2.26 (2.17, 2.35) | 5.06 (4.80, 5.34) | 1.34 (1.26, 1.42) | 4.30 (3.94, 4.69) | 2.14 (1.73, 2.66) |
| >90d | N/A | 50.61 (48.70, 52.59) | 6.60 (6.37, 6.84) | 1.21 (1.02, 1.44) | 8.02 (6.54, 9.85) | 17.38 (16.74, 18.04) | 1.50 (1.47, 1.53) | 1.09 (1.06, 1.12) | 2.34 (2.28, 2.39) | 5.22 (5.04, 5.40) | 1.55 (1.50, 1.61) | 4.92 (4.67, 5.18) | 2.97 (2.65, 3.34) |
| **Manual Therapy (MT)** | | | | | | | | | | | | | |
| 0-7d | 17.38 (17.07, | N/A | 3.46 (3.29, 3.63) | 0.73 (0.58, 0.93) | 9.15 (7.36, 11.37) | 12.86 (12.41, 13.33) | 0.70 (0.67, 0.73) | 0.82 (0.79, 0.85) | 1.39 (1.34, 1.44) | 1.66 (1.56, 1.78) | 0.64 (0.60, 0.68) | 2.11 (1.94, 2.29) | 0.78 (0.62, 0.99) |
| 8-14d | 18.91 (18.57, | N/A | 2.41 (2.22, 2.62) | 0.58 (0.40, 0.83) | 8.79 (6.53, 11.84) | 11.46 (10.90, 12.04) | 0.93 (0.89, 0.98) | 1.11 (1.06, 1.16) | 1.93 (1.86, 2.01) | 2.34 (2.17, 2.52) | 0.83 (0.77, 0.89) | 1.93 (1.72, 2.18) | 1.20 (0.92, 1.57) |
| 15-28d | 18.80 (18.47, | N/A | 2.45 (2.28, 2.63) | 0.69 (0.51, 0.92) | 6.68 (4.97, 8.97) | 11.20 (10.72, 11.71) | 1.02 (0.98, 1.06) | 1.06 (1.02, 1.10) | 2.11 (2.04, 2.17) | 2.98 (2.82, 3.16) | 0.88 (0.83, 0.93) | 2.07 (1.87, 2.29) | 1.60 (1.31, 1.95) |
| 29-60d | 18.07 (17.74, | N/A | 3.50 (3.31, 3.70) | 0.80 (0.61, 1.04) | 9.89 (7.77, 12.59) | 11.79 (11.31, 12.30) | 1.17 (1.13, 1.21) | 1.10 (1.07, 1.15) | 2.24 (2.17, 2.31) | 4.35 (4.16, 4.55) | 1.15 (1.09, 1.21) | 3.07 (2.84, 3.32) | 1.76 (1.46, 2.11) |
| 61-90d | 17.51 (17.11, | N/A | 4.74 (4.42, 5.08) | 0.92 (0.64, 1.33) | 10.44 (7.43, 14.67) | 12.38 (11.69, 13.12) | 1.32 (1.26, 1.38) | 1.11 (1.05, 1.17) | 2.15 (2.05, 2.25) | 4.63 (4.35, 4.93) | 1.28 (1.19, 1.37) | 4.09 (3.70, 4.52) | 1.74 (1.32, 2.30) |
| >90d | 17.36 (17.03, | N/A | 5.29 (5.07, 5.52) | 1.14 (0.92, 1.40) | 14.14 (11.59, 17.24) | 13.44 (12.94, 13.95) | 1.49 (1.45, 1.53) | 1.12 (1.08, 1.16) | 2.27 (2.21, 2.34) | 4.96 (4.77, 5.16) | 1.53 (1.47, 1.60) | 4.70 (4.43, 4.99) | 3.07 (2.69, 3.51) |
| **Manipulation - Chiropractic (CMT)** | | | | | | | | | | | | | |
| 0-7d | 4.37 (4.23, 4.52) | 3.87 (3.70, 4.06) | N/A | 0.42 (0.30, 0.60) | 5.33 (4.06, 6.99) | 14.30 (13.80, 14.83) | 0.69 (0.66, 0.72) | 0.89 (0.86, 0.93) | 1.40 (1.34, 1.45) | 0.92 (0.83, 1.01) | 0.79 (0.74, 0.84) | 2.09 (1.92, 2.29) | 0.85 (0.66, 1.09) |
| 8-14d | 3.88 (3.67, 4.11) | 3.44 (3.17, 3.72) | N/A | 0.67 (0.43, 1.05) | 6.57 (4.45, 9.70) | 13.41 (12.68, 14.18) | 0.87 (0.81, 0.93) | 0.92 (0.87, 0.98) | 1.60 (1.51, 1.69) | 1.20 (1.05, 1.37) | 1.04 (0.95, 1.13) | 1.65 (1.40, 1.94) | 0.90 (0.61, 1.32) |
| 15-28d | 3.76 (3.58, 3.96) | 3.28 (3.06, 3.51) | N/A | 0.60 (0.40, 0.89) | 5.37 (3.73, 7.73) | 13.24 (12.61, 13.90) | 0.95 (0.90, 1.00) | 0.85 (0.80, 0.90) | 1.31 (1.24, 1.39) | 1.24 (1.11, 1.39) | 1.05 (0.98, 1.13) | 1.40 (1.21, 1.63) | 0.81 (0.58, 1.15) |
| 29-60d | 3.69 (3.54, 3.84) | 3.42 (3.23, 3.61) | N/A | 0.47 (0.33, 0.68) | 6.16 (4.66, 8.14) | 13.07 (12.54, 13.62) | 0.96 (0.92, 1.00) | 0.75 (0.71, 0.78) | 1.31 (1.25, 1.37) | 1.18 (1.08, 1.29) | 0.98 (0.92, 1.04) | 1.31 (1.15, 1.48) | 0.89 (0.68, 1.16) |
| 61-90d | 3.41 (3.24, 3.59) | 3.04 (2.83, 3.26) | N/A | 0.49 (0.32, 0.75) | 3.19 (2.03, 5.03) | 12.89 (12.28, 13.53) | 1.01 (0.96, 1.06) | 0.72 (0.68, 0.77) | 1.16 (1.09, 1.23) | 1.10 (0.98, 1.23) | 0.98 (0.91, 1.05) | 1.37 (1.19, 1.59) | 0.72 (0.50, 1.02) |
| >90d | 4.09 (3.97, 4.23) | 3.69 (3.54, 3.86) | N/A | 0.81 (0.64, 1.01) | 6.92 (5.56, 8.61) | 13.69 (13.23, 14.16) | 1.26 (1.23, 1.30) | 0.79 (0.76, 0.82) | 1.57 (1.52, 1.62) | 1.80 (1.70, 1.91) | 1.21 (1.16, 1.26) | 2.26 (2.10, 2.44) | 1.21 (1.00, 1.46) |
| **Manipulation - Osteopathic (OMT)** | | | | | | | | | | | | | |
| 0-7d | 0.96 (0.88, 1.05) | 0.60 (0.52, 0.69) | 0.47 (0.39, 0.55) | N/A | 1.47 (0.87, 2.49) | 1.21 (1.08, 1.36) | 0.34 (0.31, 0.37) | 0.44 (0.41, 0.47) | 0.39 (0.35, 0.43) | 0.39 (0.33, 0.47) | 0.36 (0.32, 0.40) | 0.67 (0.55, 0.81) | 0.35 (0.22, 0.56) |
| 8-14d | 1.40 (0.94, 2.10) | 1.31 (0.78, 2.18) | 0.91 (0.47, 1.79) | N/A | 6.28 (1.59, 24.89) | 1.41 (0.78, 2.56) | 0.73 (0.54, 1.00) | 1.10 (0.88, 1.38) | 1.41 (1.10, 1.82) | 1.07 (0.61, 1.89) | 0.81 (0.55, 1.20) | 2.04 (1.16, 3.58) | 0.58 (0.08, 4.11) |
| 15-28d | 1.49 (1.10, 2.01) | 1.22 (0.80, 1.84) | 1.04 (0.64, 1.69) | N/A | 1.90 (0.27, 13.44) | 2.05 (1.41, 2.98) | 0.80 (0.64, 1.00) | 0.94 (0.77, 1.14) | 0.98 (0.76, 1.26) | 1.18 (0.78, 1.79) | 0.87 (0.65, 1.16) | 2.46 (1.66, 3.66) | 0.35 (0.05, 2.50) |
| 29-60d | 1.46 (1.10, 1.93) | 1.51 (1.07, 2.13) | 0.53 (0.28, 1.01) | N/A | 4.89 (1.59, 15.09) | 1.68 (1.14, 2.48) | 0.92 (0.76, 1.11) | 1.00 (0.84, 1.19) | 1.07 (0.86, 1.34) | 1.52 (1.09, 2.13) | 1.31 (1.06, 1.61) | 3.27 (2.39, 4.46) | 1.51 (0.64, 3.61) |
| 61-90d | 1.64 (1.17, 2.31) | 0.87 (0.48, 1.58) | 0.79 (0.40, 1.55) | N/A | 5.45 (1.37, 21.61) | 1.35 (0.76, 2.37) | 0.84 (0.65, 1.09) | 1.00 (0.80, 1.25) | 0.96 (0.71, 1.30) | 1.52 (0.99, 2.35) | 1.28 (0.97, 1.68) | 2.25 (1.37, 3.70) | 2.02 (0.77, 5.32) |
| >90d | 2.54 (2.14, 3.02) | 2.49 (2.00, 3.11) | 1.75 (1.31, 2.35) | N/A | 6.18 (2.59, 14.79) | 2.67 (2.05, 3.46) | 1.04 (0.90, 1.21) | 1.20 (1.05, 1.37) | 1.32 (1.12, 1.55) | 2.81 (2.30, 3.43) | 1.56 (1.33, 1.83) | 4.66 (3.75, 5.81) | 0.92 (0.35, 2.43) |
| **Acupuncture (Acu)** | | | | | | | | | | | | | |
| 0-7d | 2.93 (2.43, 3.54) | 5.35 (4.58, 6.26) | 2.67 (2.02, 3.53) | 3.42 (1.87, 6.28) | N/A | 8.22 (7.12, 9.50) | 0.47 (0.34, 0.64) | 0.60 (0.46, 0.78) | 0.90 (0.70, 1.17) | 1.36 (0.93, 1.99) | 0.65 (0.47, 0.92) | 1.71 (1.07, 2.74) | 0.34 (0.05, 2.40) |
| 8-14d | 3.24 (2.44, 4.30) | 5.62 (4.41, 7.17) | 3.72 (2.58, 5.38) | 1.82 (0.46, 7.15) | N/A | 6.33 (4.73, 8.47) | 0.88 (0.63, 1.23) | 0.72 (0.49, 1.06) | 1.33 (0.96, 1.84) | 2.71 (1.81, 4.08) | 1.13 (0.77, 1.68) | 1.99 (0.98, 4.03) | N/A |
| 15-28d | 3.66 (2.93, 4.56) | 4.71 (3.70, 6.01) | 4.18 (3.12, 5.59) | 0.67 (0.10, 4.71) | N/A | 7.39 (5.93, 9.22) | 0.78 (0.57, 1.07) | 0.90 (0.68, 1.20) | 1.21 (0.90, 1.64) | 2.22 (1.49, 3.30) | 1.14 (0.82, 1.60) | 1.88 (1.00, 3.52) | 0.66 (0.09, 4.66) |
| 29-60d | 3.52 (2.88, 4.31) | 6.46 (5.49, 7.60) | 4.85 (3.85, 6.13) | 1.59 (0.52, 4.87) | N/A | 5.72 (4.50, 7.28) | 1.09 (0.87, 1.35) | 0.80 (0.61, 1.05) | 1.36 (1.07, 1.74) | 2.99 (2.23, 4.00) | 1.04 (0.76, 1.43) | 2.81 (1.80, 4.39) | 0.52 (0.07, 3.70) |
| 61-90d | 3.69 (2.90, 4.70) | 6.46 (5.28, 7.91) | 3.67 (2.58, 5.22) | 2.45 (0.81, 7.47) | N/A | 7.07 (5.49, 9.11) | 0.99 (0.74, 1.32) | 0.75 (0.53, 1.06) | 1.24 (0.90, 1.72) | 2.44 (1.62, 3.69) | 1.67 (1.25, 2.22) | 2.55 (1.42, 4.58) | 3.24 (1.24, 8.45) |
| >90d | 4.40 (3.91, 4.96) | 6.28 (5.57, 7.08) | 5.91 (5.13, 6.82) | 2.17 (1.09, 4.29) | N/A | 6.95 (6.00, 8.07) | 1.33 (1.16, 1.51) | 0.94 (0.79, 1.11) | 1.42 (1.20, 1.69) | 3.10 (2.53, 3.80) | 1.33 (1.10, 1.62) | 3.55 (2.69, 4.69) | 1.61 (0.73, 3.55) |

Cells in red are not different than the reference of if the specific service was not performed (p=.05)
