## Supplement - State Summary for "Low back pain service utilization and costs: association with timing of first-line services for individuals initially contacting a primary care provider. A retrospective cohort study"

### Supplement - Episode count by home address state of individual with low back pain

| State | Episodes | % | | State | Episodes | % |
|---|---|---|---|---|---|---|
| Total | 347627 | 100.0% | | KS | 4040 | 1.2% |
| TX | 43028 | 12.4% | | KY | 3880 | 1.1% |
| FL | 31815 | 9.2% | | SC | 3655 | 1.1% |
| NC | 16286 | 4.7% | | OR | 3327 | 1.0% |
| OH | 15912 | 4.6% | | MA | 3104 | 0.9% |
| IL | 15661 | 4.5% | | UT | 2647 | 0.8% |
| CA | 14396 | 4.1% | | RI | 2636 | 0.8% |
| MO | 12861 | 3.7% | | CT | 2631 | 0.8% |
| WI | 12625 | 3.6% | | AL | 2108 | 0.6% |
| AZ | 12482 | 3.6% | | NV | 1754 | 0.5% |
| CO | 11814 | 3.4% | | NM | 1477 | 0.4% |
| MN | 11563 | 3.3% | | DC | 953 | 0.3% |
| MD | 10607 | 3.1% | | WV | 893 | 0.3% |
| GA | 9898 | 2.9% | | ND | 647 | 0.2% |
| VA | 9182 | 2.6% | | NH | 566 | 0.2% |
| IN | 9167 | 2.6% | | ID | 521 | 0.2% |
| TN | 8703 | 2.5% | | ME | 449 | 0.1% |
| LA | 7689 | 2.2% | | DE | 372 | 0.1% |
| NY | 7327 | 2.1% | | WY | 335 | 0.1% |
| NE | 6164 | 1.8% | | SD | 288 | 0.1% |
| AR | 5875 | 1.7% | | MT | 205 | 0.1% |
| IA | 5409 | 1.6% | | VI | 158 | 0.1% |
| MS | 5345 | 1.5% | | VT | 69 | 0.0% |
| PA | 5329 | 1.5% | | PR | 56 | 0.0% |
| OK | 5068 | 1.5% | | AK | 52 | 0.0% |
| WA | 4798 | 1.4% | | HI | 42 | 0.0% |
| MI | 4500 | 1.3% | | Unknown | 3026 | 0.9% |
| NJ | 4232 | 1.2% | | | | |
